# Hypothalamic Gliosis is Associated With Multiple Cardiovascular Disease Risk Factors

**DOI:** 10.1101/2024.09.19.24313914

**Authors:** Justin Lo, Susan J Melhorn, Sarah Kee, Kelsey LW Olerich, Alyssa Huang, Dabin Yeum, Alexa Beiser, Sudha Seshadri, Charles De Carli, Ellen A Schur

**Author notes:** Corresponding author: Ellen A Schur UW Medicine Diabetes Institute, 750 Republican Street, Box 358062, Seattle, WA 98109.

## Abstract

**Background:** Hypothalamic gliosis is mechanistically linked to obesity and insulin resistance in rodent models. We tested cross-sectional associations between radiologic measures of hypothalamic gliosis in humans and clinically relevant cardiovascular disease risk factors, as well as prevalent coronary heart disease.

**Methods:** Using brain MRI images from Framingham Heart Study participants (N=867; mean age, 55 years; 55% females), T2 signal intensities were extracted bilaterally from the region of interest in the mediobasal hypothalamus (MBH) and reference regions in the amygdala (AMY) and putamen (PUT). T2 signal ratios were created in which greater relative T2 signal intensity suggests gliosis. The primary measure compared MBH to AMY (MBH/AMY); a positive control ratio (MBH/PUT) also assessed MBH whereas a negative control (PUT/AMY) did not. Outcomes were BMI, HDL-C, LDL-C, fasting triglycerides, and the presence of hypertension (n=449), diabetes mellitus (n=66), metabolic syndrome (n=254), or coronary heart disease (n=25). Dietary risk factors for gliosis were assessed in a prospective analysis. Statistical testing was performed using linear or logistic regression.

**Results:** Greater MBH/AMY T2 signal ratios were associated with higher BMI (β = 21.5 [95% CI, 15.4– 27.6]; *P*<0.001), higher fasting triglycerides (β = 1.1 [95% CI, 0.6–1.7]; *P*<0.001), lower HDL-C (β = –20.8 [95% CI, –40.0 to –1.6]; *P*=0.034), and presence of hypertension (odds ratio, 1.2 [95% CI, 1.1–1.4]; *P*=0.0088), and the latter two were independent of BMI. Findings for diabetes mellitus were mixed and attenuated by adjusting for BMI. Metabolic syndrome was associated with MBH/AMY T2 signal ratios (odds ratio, 1.3 [95% CI, 1.1–1.6]; *P*<0.001). Model results were almost uniformly confirmed by the positive control ratios, whereas negative control ratios that did not test the MBH were unrelated to any outcomes (all *P*≥0.05). T2 signal ratios were not associated with prevalent coronary heart disease (all *P*>0.05), but confidence intervals were wide. Self-reported percentages of macronutrient intake were not consistently related to future T2 signal ratios.

**Conclusions:** Using a well-established study of cardiovascular disease development, we found evidence linking hypothalamic gliosis to multiple cardiovascular disease risk factors, even independent of adiposity. Our results highlight the need to consider neurologic mechanisms to understand and improve cardiometabolic health.

## Introduction

Obesity, hypertension (HTN), metabolic syndrome (MetS), and type 2 diabetes (T2D) are well-established risk factors for increased cardiovascular disease (CVD) morbidity and mortality. However, central nervous system (CNS) involvement in the pathogenesis of CVD risk factors, including obesity and T2D, is increasingly recognized.^1^ A robust preclinical literature demonstrates that diet-induced obesity in animal models is causally related to cellular inflammatory responses in the hypothalamus.^2^ Furthermore, T2D is linked to the remodeling of extracellular matrix structures that scaffold neurons and their synaptic connections.^3^ In humans, *ex vivo* histopathological and *in vivo* neuroimaging studies support that cellular and tissue-level alterations consistent with hypothalamic gliosis are present in the human hypothalamus in association with obesity and metabolic disease.^1^ Prospective studies further support a role in obesity and T2D pathogenesis by showing that the extent of gliosis, defined as proliferation and inflammatory activation of glia, is associated with weight gain and progressive insulin resistance in children and adults with overweight or obesity.^4,5^ Moreover, arcuate nucleus inflammation may mediate the increased atherogenic risk associated with hypogonadism in males.^6^ Further studies have linked hypothalamic inflammation to MetS, T2D, and impaired glucose tolerance.^5,7,8^ Therefore, hypothalamic inflammation and gliosis could indirectly increase the risk of CVD by promoting metabolic dysregulation, chronic systemic inflammation, and/or atherogenic lipid profiles.

Alternatively, diet-induced hypothalamic inflammation in the MBH and its arcuate nucleus could directly contribute to CVD development. The arcuate nucleus receives sensory inputs and transmits intra- and extra-hypothalamic communications to govern homeostatic regulation of key metabolic functions including energy balance, feeding behavior, and maintenance of circulating glucose, but also sympathetic nerve activity (SNA).^9^ The latter involves reciprocal inputs, primarily via the paraventricular nucleus, that can increase sympathetic tone, peripheral vascular resistance, and, hence, blood pressure.^10^ Based on these delineated neuronal pathways, inflammation and gliosis affecting the arcuate nucleus has the potential to underlie obesity-associated increases in blood pressure and prevalence of HTN. This has yet to be investigated and offers a route, separate from the above-noted metabolic conditions, whereby CVD risk could be elevated via a CNS etiology. In summary, the accumulated literature suggests hypothalamic inflammation and gliosis could influence CVD development. We therefore investigated, in the Framingham Heart Study (FHS), the associations of multiple cardiometabolic risk factors as well as prevalent coronary heart disease (CHD) with radiologically assessed measures of hypothalamic gliosis. We also prospectively examined dietary risk factors for hypothalamic gliosis based on self-reported dietary intake.

## Methods

All data used in the analyses were shared through the FHS Repository and are available by request for approved projects. JL had full access to the data and is responsible for the integrity of the data analysis in the study.

### Study population

In this cross-sectional study, 905 MRI images were provided from magnetic resonance imaging (MRI) exams conducted on the FHS Offspring and Generation 3 cohorts (Figure S1). The FHS is a longitudinal, multigenerational observational cohort study investigating cardiovascular risk factors. FHS cohort recruitment and enrollment are described in detail elsewhere.^11,12^ Brain MRI was added to the FHS to investigate brain aging.^13^ We evaluated 3T MRI images assessed between 2012 and 2018 in relation to cardiovascular health indicators measured in the Offspring (examination 9, 2011–2014, n=25) and Generation 3 (examination 2, 2009–2011, n=14; examination 3, 2016–2019, n=828) cohorts. For the majority of the sample, brain MR imaging was conducted in proximity to examination 3, on average, –0.3 years (range, –3.1 to 2.3 years) before examination 3 for Generation 3 cohort participants. For the remaining participants, the MRI occurred an average of 5.6 years (range, 2.5–9.2 years) after examination 9 for Offspring Cohort participants and 6.4 years (range, 5.9–7.1 years) after examination 2 for Generation 3 participants without examination 3 data. 35 MRI images were uninterpretable in regions of interest (ROIs), while 3 MRI images were repeated scans from the same individual. The total sample size included 867 individuals with unique MRI images and ratable T2-signal intensities in ROIs. Those with missing data for certain covariates or outcomes were excluded from relevant sub-analyses. For triglyceride analyses, we excluded individuals with fasting triglyceride concentrations above 500 mg/dL (n=4).

### Outcomes of interest

Outcomes of interest in this study were cardiovascular risk factors including body mass index (BMI), high-density lipoprotein cholesterol (HDL-C), low-density lipoprotein cholesterol (LDL-C), fasting triglycerides, HTN, diabetes mellitus (DM), and MetS, as well as prevalent coronary heart disease (CHD). BMI was calculated from height and weight measurements. HDL-C, LDL-C, and fasting (at least 8 hours prior to blood sample collection) triglycerides were measured in ethylenediaminetetraacetic acid serum tests. The presence of HTN was based on the American Heart Association stage 1 and stage 2 HTN definitions as either systolic blood pressure greater than or equal to 130 mmHg, diastolic blood pressure greater than or equal to 80 mmHg, and/or receiving treatment for HTN.^14^ DM was defined as either fasting blood glucose greater than or equal to 126 mg/dL, random blood glucose greater than or equal to 200 mg/dL, and/or treatment for DM. Distinction between type 1 diabetes (T1D) and T2D was not ascertained but most cases were assumed to be T2D, as discussed in previous literature.^15^ MetS was defined as having 3 or more criteria of metabolic abnormalities according to the American Heart Association and the National Heart, Lung, and Blood Institute definition^16^: 1) systolic blood pressure greater than or equal to 130 mmHg, diastolic blood pressure greater than or equal to 85 mmHg, or receiving treatment for HTN; 2) fasting blood glucose greater than or equal to 100 mg/dL or receiving treatment for elevated blood glucose; 3) HDL-C less than 40 mg/dL in males or 50 mg/dL in females or receiving treatment for low HDL-C; 4) triglycerides greater than or equal to 150 mg/dL or receiving treatment for high triglycerides; 5) waist circumference greater than or equal to 102 cm for males or 88 cm for females. Finally, CHD was defined as manifestations of CHD before or during the FHS study period, including myocardial infarction, coronary insufficiency, angina pectoris, sudden death from CHD, and non-sudden death from CHD, and was adjudicated by FHS investigators.

### Dietary Exposures

In a prospective cohort analysis, the relation of dietary exposures to subsequent MRI findings was tested. Dietary exposures were collected from a Food Frequency Questionnaire (FFQ) administered at the FHS exam prior to MRI acquisition (Offspring cohort, examination 8; Generation 3 cohort, examination 2). Daily consumptions of the proportions of total fat, total carbohydrates, total protein, total sugar, fructose, sucrose, and saturated fat of total energy intake were calculated by dividing each dietary exposure intake (kilocalories consumed) by the average daily caloric intake. 51 participants were missing FFQ data and 2 participants with implausible values for the proportions of total fat and saturated fat were removed from all dietary analyses, leaving a total sample of N=814. There was a greater percentage of females in the group of participants with FFQ data compared to those without; otherwise, the two groups did not differ in other relevant characteristics such as age, smoking status, and BMI (data not shown).

### Covariates

Age, sex, and smoking status were collected and reported in the FHS exam data. Information on an individual’s treatment status for HTN, DM, and lipid levels was determined from both self-reported exam data and medication review. Physical activity was captured based on a physical activity index derived from self-reported levels of activity during a typical 24-hour period. Methods to construct the physical activity index are described in detail elsewhere.^17^

### MRI Acquisition and Analysis

High-resolution T2-weighted axial brain images were analyzed in Horos Medical Image Software by two independent raters (Figure 1). The image slice superior to the optic chiasm, with visible optic tracts and mammillary bodies, was identified as the MBH slice of interest. The MBH ROI was placed bilaterally in the anterior hypothalamic area adjacent to the 3rd ventricle to encompass the region of the arcuate nucleus. Slices inferior and superior were selected based on clear anatomical landmarks for the control ROIs (i.e., AMY and PUT). T2 signal intensities were extracted bilaterally from the regions (i.e., MBH, AMY, and PUT). The signal intensities were then used to create ratios for the left, right, and bilateral mean for our primary measure (MBH/AMY) T2 signal ratio assessing hypothalamic gliosis, a positive control (MBH/PUT) T2 signal ratio, and a negative control (PUT/AMY) T2 signal ratio. T2 signal ratios were used to capture MBH T2 signal relative to gray matter reference ROIs and to account for inter-subject variability in signal intensity of the T2-weighted acquisitions. Calculated ratios from individual raters were averaged and used in statistical analyses. The intraclass correlations between the independent raters for the MBH/AMY, MBH/PUT, and PUT/AMY ratios were 0.71, 0.82, and 0.91, respectively.

**Figure 1.**
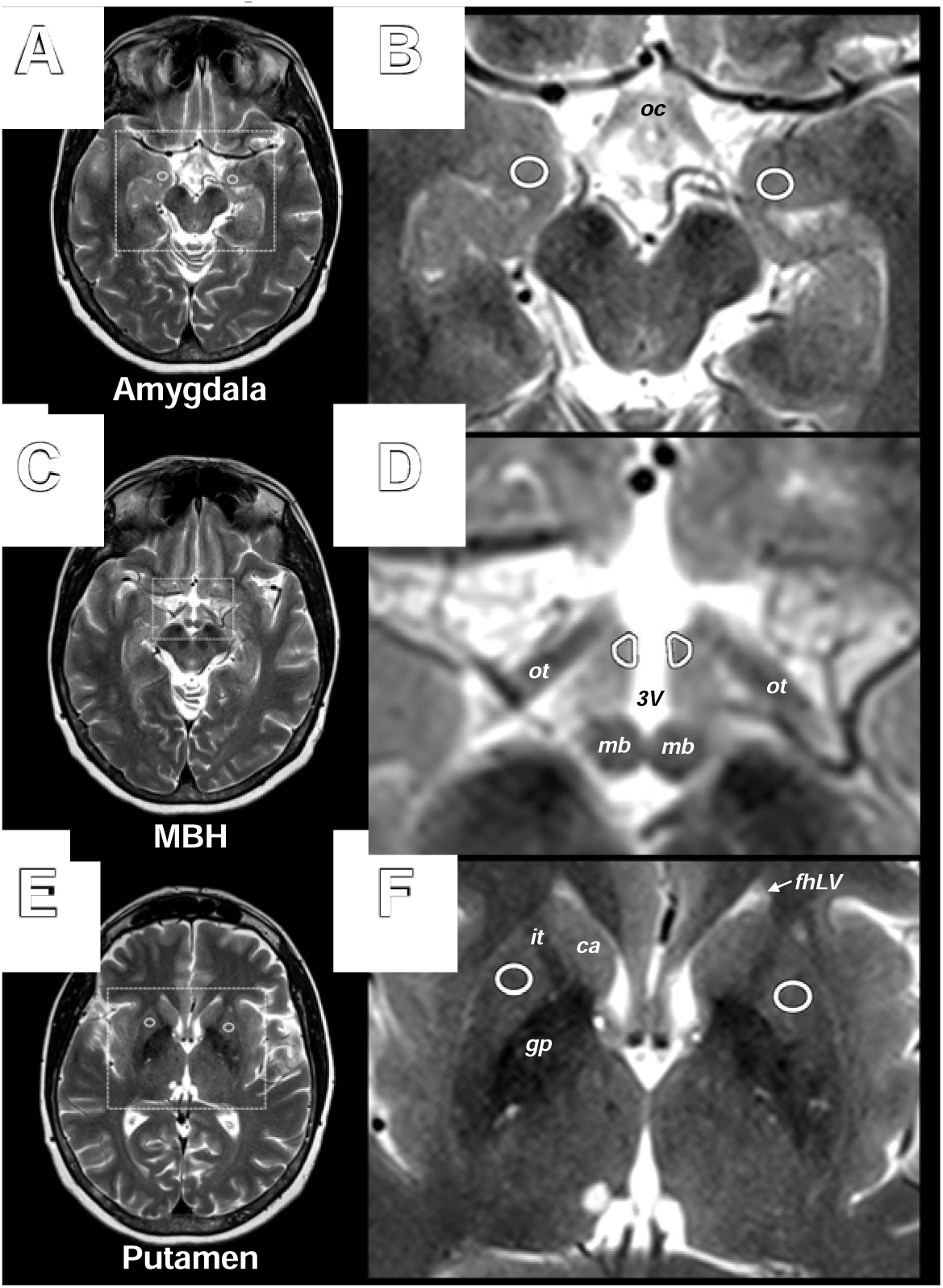
Axial brain T2-weighted image with anatomic locations of MBH and reference regions of interest. Representative MRI images and regions of interest (ROIs). A) The amygdala was identified at the level of the optic chiasm (oc) which was, on average, one slice inferior to the hypothalamus. B) Inset identified in A with representative bilateral placement of ROIs in the amygdala. C) The mediobasal hypothalamus (MBH) was identified on the slice with distinct optic tracts (ot) and visible mammillary bodies (mb) and is at the level of the superior colliculus of the midbrain (superior to A). D) Inset identified in C with representative placement of bilateral ROIs in the anterior hypothalamus, encompassing the location of the arcuate nucleus, adjacent to the 3^rd^ ventricle. E) The putamen was identified at a level of the frontal horn of the lateral ventricles (with visible caudate head, internal capsule and globus pallidus). F) Inset identified in E with representative placement of bilateral ROIs in the putamen. oc, optic chiasm; MBH, mediobasal hypothalamus; ot, optic tract; 3V, 3^rd^ ventricle; mb, mammillary body; fhLV, frontal horn of the lateral ventricle; ca, caudate; it, internal capsule; gp, globus pallidus.

### Statistical analysis

Multiple linear regression was used to analyze continuous outcomes (i.e., BMI, LDL-C, HDL-C, and triglycerides); multiple logistic regression was used to analyze binary outcomes (i.e., HTN, DM, MetS, and CHD). Coefficients and confidence intervals are reported as the estimated change in outcome per 1 unit difference in the predictor variable, while odds ratios and confidence intervals are reported as the estimated change in odds per 1 SD difference in the predictor variable. Fasting triglyceride levels were right-skewed and natural logarithm-transformed for analysis. T2 signal ratios for the primary exposure, positive control, and negative control predictors were natural logarithm transformed and used as predictors in the models to satisfy conditions of normality. Age and sex were included as covariates in the base model (Model 1). Current smoking status was added in subsequent models (Model 2), while risk factor treatment status was further adjusted for outcomes except MetS (Model 3). BMI was added in fully adjusted models when appropriate. Table S1 describes the full set of covariates included in the models. We evaluated statistical significance using two-sided tests at the *P*<0.05 level, uncorrected for multiple comparisons for our *a prior*i outcomes, to test associations with and without adjustment for confounders such as BMI. Statistics were computed in R version 4.2.3.

To test associations between dietary exposures and T2 signal ratios, both simple and multiple linear regression models were conducted adjusting for age, sex, and time interval between dietary exposure measurement and MRI measurement (years). A false discovery rate (FDR) multiple comparison correction was applied to this set of analyses at *q*<0.05. To explore if the associations were modified by weight status, interaction terms between BMI categories (<25 kg/m^2^; ≥25 kg/m^2^ & <30 kg/m^2^; ≥30 kg/m^2^) and each dietary exposure were added in separate regression models. Diet analysis was conducted using R version 4.0.2.

## Results

### Participant characteristics

Characteristics from 867 individuals are presented in Table 1. The sample averaged 55 years of age and was 55% female. The mean BMI was in the overweight range, whereas 34.6% of the sample had a BMI in the obesity range. Current smoking (5.4%), diabetes treatment (5.4%), and prevalent CHD (2.9%) were rare.

**Table 1.** Study population characteristics.

|  | Summary (N=867) |
| --- | --- |
| Age, years, mean (SD) | 54.9 (8.8) |
| Sex |  |
| Female, n (%) | 476 (54.9) |
| Male, n (%) | 391 (45.1) |
| Self-reported race |  |
| White, n (%) | 856 (98.7) |
| Black, n (%) | 1 (0.1) |
| Other, n (%) | 10 (1.2) |
| Smoking status* |  |
| Does not smoke, n (%) | 819 (94.6) |
| Currently smokes, n (%) | 47 (5.4) |
| FHS physical activity index, mean (SD)<sup>†</sup> | 36.0 (6.2) |
| Body mass index, kg/m<sup>2</sup>, mean (SD) | 28.6 (5.5) |
| BMI<25 kg/m<sup>2</sup>, n (%) | 240 (27.7) |
| 25≤BMI<30 kg/m<sup>2</sup>, n (%) | 327 (37.7) |
| BMI≥30 kg/m<sup>2</sup>, n (%) | 300 (34.6) |
| Waist circumference, cm, mean (SD) | 99.7 (14.4) |
| HDL cholesterol, mg/dL, mean (SD)<sup>‡</sup> | 60.2 (20.0) |
| LDL cholesterol, mg/dL, mean (SD)<sup>§</sup> | 106.3 (29.4) |
| Fasting triglycerides, mg/dL, mean (SD)<sup>‡, \|\|, #</sup> | 111.8 (72.0) |
| Lipid treatment |  |
| Not treated for lipids, n (%) | 638 (73.6) |
| Treated for lipids, n (%) | 229 (26.4) |
| Systolic blood pressure, mm Hg, mean (SD) | 119.6 (14.0) |
| Diastolic blood pressure, mm Hg, mean (SD)** | 76.0 (8.8) |
| Hypertension treatment*** |  |
| Not treated for hypertension, n (%) | 641 (74.0) |
| Treated for hypertension, n (%) | 225 (26.0) |
| Hypertension (HTN) |  |
| Absence of HTN, n (%) | 417 (48.2) |
| Prevalent HTN, n (%) | 449 (51.8) |
| Blood glucose, mg/dL, mean (SD)<sup>‡</sup> | 99.6 (20.0) |
| Diabetes mellitus treatment |  |
| Not treated for diabetes, n (%) | 820 (94.6) |
| Treated for diabetes, n (%) | 47 (5.4) |
| Diabetes mellitus (DM)<sup>‡</sup> |  |
| Absence of DM, n (%) | 798 (92.4) |
| Prevalent DM, n (%) | 66 (7.6) |
| Metabolic syndrome (MetS) |  |
| Absence of MetS, n (%) | 613 (70.7) |
| Prevalent MetS, n (%) | 254 (29.3) |
| Coronary heart disease (CHD) |  |
| Absence of CHD, n (%) | 842 (97.1) |
| Prevalent CHD, n (%) | 25 (2.9) |
| MBH/AMY T2 signal ratio, mean (SD) | 1.27 (0.07) |
| MBH/PUT T2 signal ratio, mean (SD)**** | 1.72 (0.14) |
| PUT/AMY T2 signal ratio, mean (SD)**** | 0.74 (0.06) |

\*n=1 individual missing information about smoking status; †n=3 individuals missing information about physical activity; ‡ n=2 individuals missing information on triglycerides, HDL cholesterol, blood glucose levels, and diabetes mellitus status; § n=12 individuals missing information on LDL cholesterol; || n=3 individuals excluded with fasting triglyceride concentrations greater than 500 mg/dL; # n=20 individuals missing fasting triglyceride concentrations; \*\*n=1 individual missing information on DBP; \*\*\* n=1 individual missing information on hypertension treatment; \*\*\*\* n=14 individuals missing MBH/PUT & PUT/AMY T2 signal ratios.

### Body mass index (BMI)

MBH/AMY T2 signal ratios and BMI were positively associated (Figure 2A). The positive association between MBH/AMY T2 signal ratios and BMI persisted after adjusting for age, sex, smoking status, and diabetes treatment (Table 2). Coefficients estimate that a 0.1 unit increase in the natural logarithm-transformed MBH/AMY T2 signal ratio would be associated with a 2.2 kg/m^2^ (95% CI 1.5–2.8) higher BMI. The positive control (MBH/PUT) T2 signal ratio was also positively associated with BMI in all models, whereas there were null associations for the negative control (PUT/AMY) T2 signal ratio and BMI (Table 2).

**Figure 2.**
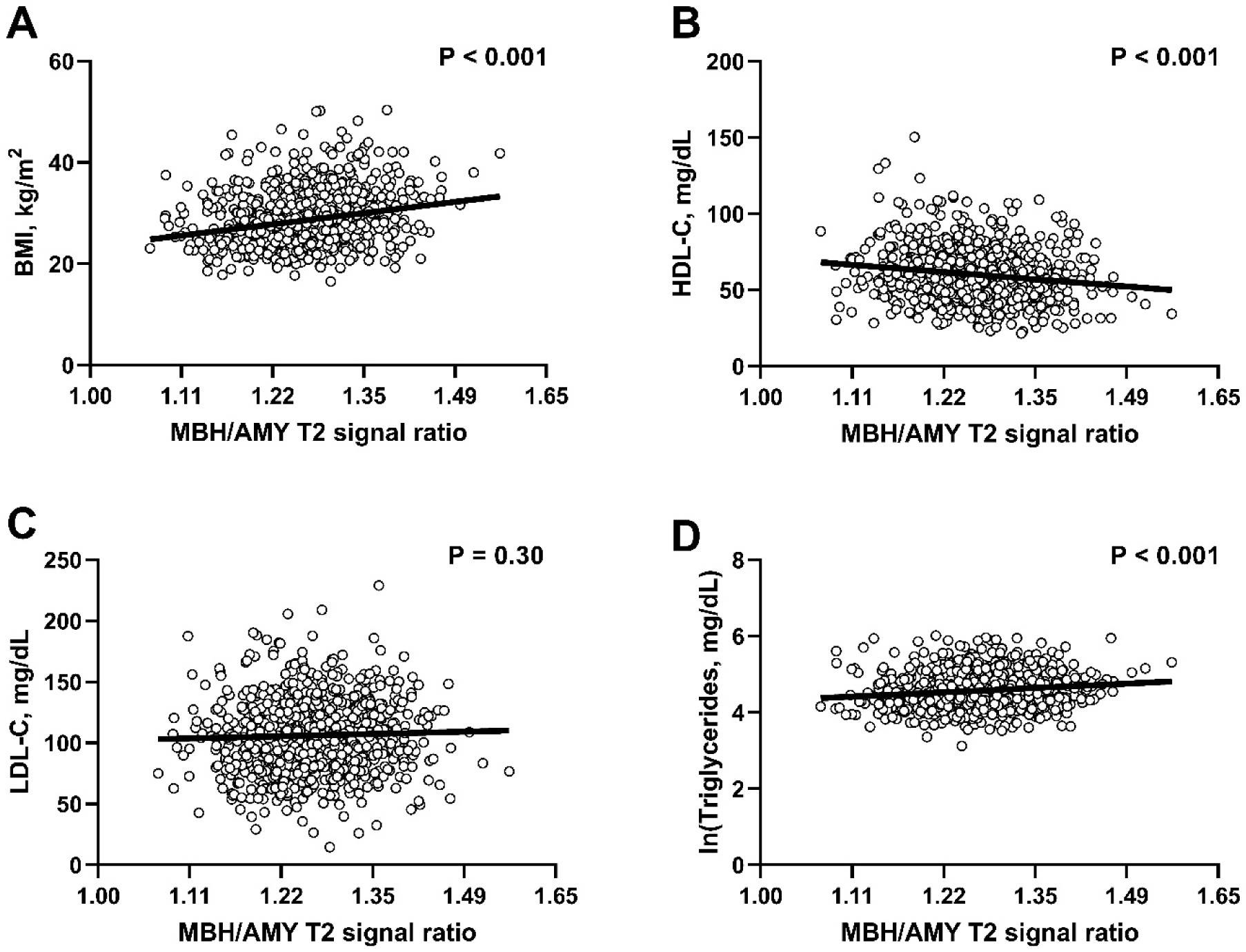
Sex- and age-adjusted associations of CVD risk factors with radiologic evidence of hypothalamic gliosis by MBH/AMY T2 signal ratios. Scatterplots with linear fit lines of adjusted associations between MBH/AMY T2 signal ratio and body mass index (BMI, A), HDL cholesterol (HDL-C, B), LDL cholesterol (LDL-C, C), and natural logarithm transformed fasting triglycerides (D). X-axis values represent MBH/AMY T2 signal ratios back-transformed from their natural logarithm value in the distribution derived from the linear fit performed on the natural logarithm transformed predictor variable. P-values correspond to the two-sided probability of the estimated t-statistic for the coefficient of the predictor in Model 1. MBH, mediobasal hypothalamus; AMY, amygdala.

**Table 2.** Associations of quantitative MRI measures and continuous outcomes related to cardiovascular risk.

| T2 signal ratio predictor and model |  | BMI |  | HDL-C |  | LDL-C |  | ln(Triglycerides) |  |
| --- | --- | --- | --- | --- | --- | --- | --- | --- | --- |
|  |  | Coefficient<br>[95% CI] | P<br>value | Coefficient<br>[95%CI] | P<br>value | Coefficient<br>[95%CI] | P<br>value | Coefficient<br>[95%CI] | P<br>value |
| MBH/AMY | Model 1* | 22.2 [16.0, 28.3] | <0.001 | -47.3 [-67.5, -27.1] | <0.001 | 18.2 [-16.5, 52.8] | 0.30 | 1.2 [0.6, 1.7] | <0.001 |
|  | Model 2† | 22.2 [16.0, 28.4] | <0.001 | -47.3 [-67.4, -27.1] | <0.001 | 17.7 [-16.9, 52.4] | 0.32 | 1.2 [0.6, 1.7] | <0.001 |
|  | Model 3‡ | 21.5 [15.4, 27.6] | <0.001 | -46.3 [-66.2, -26.3] | <0.001 | 21.5 [-11.3, 54.4] | 0.20 | 1.1 [0.6, 1.7] | <0.001 |
|  | Fully adjusted§ | ... | ... | -20.8 [-40.0, -1.6] | 0.034 | 12.5 [-21.2, 46.3] | 0.47 | 0.5 [-0.0, 1.0] | 0.076 |
| MBH/PUT | Model 1* | 12.7 [8.3, 17.0] | <0.001 | -34.2 [-48.3, -20.2] | <0.001 | -2.0 [-26.0, 22.1] | 0.87 | 0.9 [0.5, 1.3] | <0.001 |
|  | Model 2† | 12.9 [8.5, 17.2] | <0.001 | -33.6 [-47.7, -19.5] | <0.001 | -1.1 [-25.2, 23.0] | 0.93 | 0.9 [0.5, 1.2] | <0.001 |
|  | Model 3‡ | 12.2 [7.9, 16.5] | <0.001 | -32.3 [-46.3, -18.4] | <0.001 | 3.0 [-19.9, 25.9] | 0.80 | 0.8 [0.4, 1.2] | <0.001 |
|  | Fully adjusted§ | ... | ... | -17.9 [-31.2, -4.6] | 0.0083 | -3.0 [-26.3, 20.2] | 0.80 | 0.5 [0.1, 0.8] | 0.012 |
| PUT/AMY | Model 1* | -2.5 [-7.4, 2.4] | 0.32 | 14.5 [-1.3, 30.3] | 0.072 | 17.4 [-9.2, 44.0] | 0.20 | -0.4 [-0.8, 0.1] | 0.085 |
|  | Model 2† | -2.7 [-7.7, 2.2] | 0.28 | 13.6 [-2.2, 29.5] | 0.092 | 16.2 [-10.5, 42.9] | 0.24 | -0.3 [-0.8, 0.1] | 0.11 |
|  | Model 3‡ | -2.2 [-7.1, 2.6] | 0.37 | 12.4 [-3.3, 28.1] | 0.12 | 12.7 [-12.7, 38.2] | 0.33 | -0.3 [-0.8, 0.1] | 0.13 |
|  | Fully adjusted§ | ... | ... | 9.7 [-4.8, 24.3] | 0.19 | 13.7 [-11.7, 39.0] | 0.29 | -0.3 [-0.7, 0.1] | 0.20 |

Results are from multiple linear regression models. MRI-assessed T2 signal ratios were natural logarithm transformed and used as model predictors: MBH/AMY (primary), MBH/PUT (positive control), and PUT/AMY (negative control). Coefficient and confidence intervals represent the estimated change in outcome per 1 unit difference in the log-transformed T2 signal ratio.
BMI, body mass index; HDL-C, high-density lipoprotein cholesterol; LDL-C, low-density lipoprotein cholesterol; ln(Triglycerides), natural logarithm transformed fasting triglycerides; MBH, mediobasal hypothalamus; AMY, amygdala; PUT, putamen.
\* Model 1 adjusted for age and sex.
† Model 2 adjusted for model 1 covariates plus smoking.
‡ Model 3 adjusted for model 2 covariates plus diabetes treatment for BMI model or lipid treatment for HDL-C, LDL-C, and natural log-transformed triglycerides models.
§ Fully adjusted model includes model 3 covariates plus BMI, when appropriate.

### HDL cholesterol (HDL-C)

There was a negative association between MBH/AMY T2 signal ratios and HDL-C (Figure 2B). Negative associations were present at all levels of adjustment in models testing the primary exposure (MBH/AMY T2 signal ratio) and the positive control (MBH/PUT) T2 signal ratio (Table 2), including after adjustment for BMI. PUT/AMY T2 signal ratios were not associated with HDL-C concentrations (Table 2).

### LDL cholesterol (LDL-C)

The primary exposure T2 signal ratios were not associated with LDL-C in model 1 (Figure 2C) nor any further adjusted models (Table 2). The positive and negative control T2 signal ratios were similarly not associated with LDL-C (Table 2).

### Triglycerides

There were positive associations between the MBH/AMY T2 signal ratios and fasting triglycerides in Model 1 (Figure 2D), as well as Models 2 and 3 (Table 2). However, the magnitude of the association was reduced such that only a trend persisted after further adjusting for BMI in the fully adjusted model (Table 2, *P*=0.076). In contrast, MBH/PUT T2 signal ratios were uniformly positively associated with natural logarithm-transformed fasting triglycerides, including after adjustment for BMI. PUT/AMY T2 signal ratios were not significantly associated with fasting triglycerides (Table 2).

### Hypertension

For all models, there were positive associations between MBH/AMY T2 signal ratios and the presence of HTN (Figure 3). Model 1 estimates that a 1 SD difference in the natural-logarithm transformed MBH/AMY T2 signal ratio is associated with a 1.4 (95% CI 1.2–1.6) times greater odds of HTN diagnosis. Positive control T2 signal ratios were also positively associated with the presence of HTN, except for in the fully adjusted model (*P*=0.12). No associations were found with the negative control T2 signal ratio (Figure 3).

**Figure 3.**
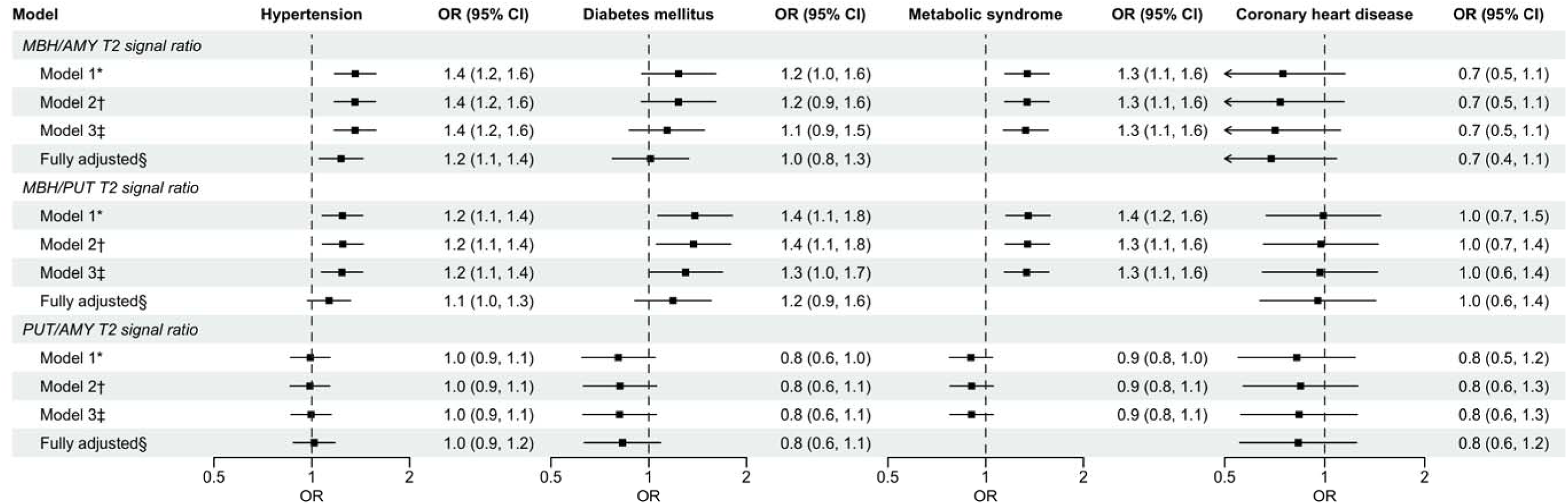
Forest plot of odds ratios & 95% confidence intervals for T2 signal ratios and binary outcomes related to cardiovascular risk and coronary heart disease. Results are from multiple logistic regression models. MRI-assessed T2 signal ratios were natural logarithm transformed and used as model predictors: MBH/AMY (primary), MBH/PUT (positive control), and PUT/AMY (negative control). Black squares represent the point estimates of the ORs for each model’s predictor, and solid lines represent 95% CIs for the ORs. ORs and 95% CIs are presented as the change in odds for the outcome per a 1 SD difference in natural logarithm-transformed T2 signal ratio. Arrows indicate that the width of the 95% CI lies outside of the figure axis. The dashed vertical line signifies the null OR of 1. BMI, body mass index; HDL-C, high-density lipoprotein cholesterol; LDL-C, low-density lipoprotein cholesterol; ln(Triglycerides), natural logarithm transformed fasting triglycerides; MBH, mediobasal hypothalamus; AMY, amygdala; PUT, putamen. * Model 1 adjusted for age and sex. † Model 2 adjusted for model 1 covariates plus smoking. ‡ Model 3 adjusted for model 2 covariates plus diabetes treatment for BMI model or lipid treatment for HDL-C, LDL-C, and natural log-transformed triglycerides models. § Fully adjusted model includes model 3 covariates plus BMI, when appropriate.

### Diabetes mellitus (DM)

MBH/AMY T2 signal ratios were not associated with prevalent DM in any models (Figure 3). MBH/PUT T2 signal ratios were associated with higher odds of prevalent DM (Figure 3), but not in the fully adjusted model that included BMI (*P*=0.21). PUT/AMY T2 signal ratios were not associated with DM (Figure 3).

### Metabolic syndrome (MetS)

MBH/AMY T2 signal ratios were positively associated with prevalent MetS in all models (Figure 3), including after adjustment for age, sex, smoking status, and physical activity. The MBH/PUT T2 signal ratios were also positively associated with prevalent MetS at all levels of adjustment, whereas the PUT/AMY T2 signal ratios were not associated (Figure 3).

### Coronary heart disease (CHD)

Among 25 individuals with prevalent CHD, 20 individuals were diagnosed with CHD prior to their MRI exam. There were no associations between the MBH/AMY T2 signal ratios and prevalent CHD (Figure 3), and all models returned coefficients with wide confidence intervals. Neither positive nor negative control T2 signal ratios were associated with prevalent CHD (Figure 3).

### Sensitivity analyses

The timing of health information collection and MRI assessment across the sample varied by individual and cohort. Results adjusted for time intervals between FHS health examination and MRI examination are presented in Tables S3 and S4, and findings were unchanged from the primary analysis.

### Dietary exposures in relation to MBH gliosis

In analyses adjusting for age, sex, and time interval between dietary exposures and MRI, no statistically significant prospective associations (Table S5) were found between self-reported intake of total fat (β=0.04 [95% CI, –0.02 to 0.11]; *P*=0.19), total carbohydrates (β=–0.04 [95% CI, –0.09 to 0.01]; *P*=0.15), total protein (β = 0.09 [95% CI, –0.04 to 0.22]; *P*=0.19), fructose (β = –0.03 [95% CI, –0.22 to 0.17]; *P*=0.78), sucrose (β = –0.08 [95% CI, –0.20 to 0.04]; *P*=0.19), or saturated fat (β=0.12 [95% CI, –0.04 to 0.29]; *P*=0.14) and the degree of MBH gliosis (natural logarithm transformed MBH/AMY T2 signal ratio) assessed at MRI examinations conducted, on average, 7 years after completion of FFQs. Negative associations were found between the proportion of daily total carbohydrate intake (β=–0.08 [95% CI, –0.16 to 0.00]; *P*=0.04) and sucrose intake (β=–0.20 [95% CI, –0.38 to –0.02]; *P*=0.03) and the positive control (MBH/PUT) T2 signal ratio. These associations did not meet the FDR correction criteria at *q*<0.05. No associations were found between dietary exposures and the negative control (PUT/AMY) T2 signal ratios (Table S5). Exploratory analyses demonstrated a significant interaction between the BMI group and the proportion of daily saturated fat intake on the degree of MBH gliosis (natural logarithm transformed MBH/AMY T2 signal ratio) after covariate adjustment (β-int=0.25 [95% CI, 0.04–0.46]; *P*-int=0.02). As shown in Figure S2, stratified analyses revealed opposing directions of slopes by BMI group (BMI<25 kg/m^2^: β=–0.31 [95% CI, –0.60 to –0.15]; *P*=0.04; 25 kg/m^2^≤BMI<30 kg/m^2^: β=0.16 [95% CI, –0.08 to 0.40]; *P*=0.20; BMI ≥30 kg/m^2^: β=0.22 [95% CI, –0.08 to 0.53]; *P*=0.14).

## Discussion

Using a well-established study of CVD development, we found evidence linking hypothalamic gliosis to multiple CVD risk factors. Prior human studies indicate that obesity and metabolic dysregulation, including glucose intolerance and T2D, are associated with histologic and radiologic signs of MBH inflammation and gliosis.^1^ In the largest sample studied to date, we found strong positive associations of MBH gliosis with BMI and MetS, supporting previous findings.^7,18^ Novel results linked MBH gliosis to low HDL-C and increased prevalence of HTN. These associations persisted with adjustment for BMI, suggesting that it is not solely obesity-related metabolic dysfunction driving the result, but that the function of central pathways regulating circulating lipids and blood pressure could be directly impacted by glial cell inflammatory responses in the MBH. Although we did not find associations between hypothalamic gliosis and a clinical diagnosis of CHD, the sample size was small, and incident disease was not assessed. In sum, these findings add to an expanding body of evidence demonstrating the influence of CNS factors, namely hypothalamic inflammation and gliosis, on the pathogenesis of cardiometabolic risk and, potentially, CVD itself.

Low levels of circulating HDL-C and high levels of LDL-C and triglycerides confer an increased risk for CVD development and are strongly associated with obesity.^19,20^ However, few studies have closely examined the relationship of dyslipidemia to diet-induced hypothalamic inflammation. A study of 41 healthy adult males found that longer MBH T2 relaxation times were associated with higher LDL-C levels but not associated with HDL-C or triglyceride levels.^6^ In a cross-sectional study, Kreutzer et al. found no association between unilateral left-sided MBH/AMY T2 signal ratios and triglyceride levels among 111 adults with and without obesity.^21^ Contrary to these smaller studies, we observed a strong negative association between MBH gliosis and HDL-C. The negative association was present even after adjusting for BMI, a finding that is consistent with the hypothesis that the severity of hypothalamic inflammation is directly related to the worsening of obesity-associated metabolic dysregulation of both peripheral glucose and blood lipids. Apolipoprotein A-I deficiency in mice produces a phenotype that recapitulates many features of MetS including increased adiposity, visceral and hepatic fat deposition, hypertriglyceridemia, glucose intolerance, and low HDL-C and is accompanied by inflammation and reactive astrocytosis in the MBH.^22^ Intriguingly, administering reconstituted human HDL-C normalized values of astrocyte inflammatory markers, indicating a potentially protective role for HDL-C in reducing hypothalamic inflammation with concomitant improvement in some features of MetS.^22^ On the contrary, randomized trials of pharmacotherapies to increase HDL-C levels have not been shown to decrease CVD risk in humans.^23–25^ Nonetheless, given the strong relationships between low HDL-C and altered hypothalamic tissue structure in the current study, hypothalamic gliosis represents a novel, largely unexplored mechanism to advance understanding of both the etiology of low HDL-C in obesity and contradictory findings regarding the benefits of raising HDL-C for reducing CVD risk. We also found evidence of a positive association between MBH/AMY T2 signal ratios and fasting triglyceride concentrations. The association was attenuated when adjusting for BMI, indicating that high levels of peripheral adiposity at least partially explain the relationship. Lastly, we did not observe associations between T2 signal ratios and LDL-C. Nevertheless, the MBH’s role in energy homeostasis and the current findings should prompt further investigation into dysregulation of circulating lipids by hypothalamic inflammation.

Preclinical evidence points to the role of the arcuate nucleus in altering blood pressure through SNA.^10,26^ High-fat diets and obesity activate the arcuate nucleus, located within the MBH, to alter SNA primarily through insulin and leptin signaling.^10,26^ Both hormones bind to pro-opiomelanocortin (POMC) and agouti-related peptide (AgRP) neurons in the arcuate nucleus, which in turn relay signals to other hypothalamic sites, such as the paraventricular nucleus (PVN), an important regulator of SNA and blood pressure.^27,28^ Specifically, POMC neurons release α–melanocyte-stimulating hormone that activates downstream melanocortin-4 receptors to increase SNA in the PVN, while AgRP neurons release neuropeptide Y (NPY) that suppresses sympathoinhibitory effects in the PVN.^29–33^ Our results provide novel evidence that MBH gliosis, a diet-induced phenomenon, is associated with HTN in humans. While the exact mechanisms remain uncertain, hyperleptinemia, hyperinsulinemia, and/or local inflammation itself may exacerbate arcuate nucleus-induced SNA, thereby raising blood pressure via the well-defined neural circuitry outlined above. In the current study, associations were attenuated but still present after adjusting for BMI, providing further rationale for investigating how inflammation and gliosis in the arcuate nucleus region could evoke adiposity-independent effects on neural circuits controlling blood pressure.

Hypothalamic gliosis has been linked to the development and progression of insulin resistance. In a recent study in humans, Rosenbaum et al. showed the degree of MBH gliosis, quantitatively assessed by MRI, was greater in individuals with obesity and impaired glucose tolerance or T2D.^5^ This result was also observed in a separate study of women with obesity and T2D.^8^ Following study subjects over time, Rosenbaum et al. found that MBH gliosis was related to worsening insulin sensitivity over a 1-year follow-up period.^5^ Our current study did not consistently detect an association between MBH T2 signal ratios and the presence of DM— however, the point estimates trended in the positive direction but wide confidence intervals made the association nonsignificant. The inability to distinguish between individuals with T1D and T2D and the relatively low prevalence of DM in this FHS sample may have limited the ability to detect a significant association. Studies assessing incident disease are needed to investigate the role that MBH gliosis might play in T2D pathogenesis.

MetS was strongly associated with greater evidence of MBH gliosis. This is perhaps unsurprising given that it is a composite clinical definition that includes CVD risk factors that were individually associated with MBH gliosis. Kullman et al. previously demonstrated that individuals with MetS had higher water content in regions of the brain including the hypothalamus, thalamus, and fornix. Water content in these regions, as suggested by the authors, may have resulted from an inflammatory response by glial cells increasing water uptake, potentially damaging local microenvironments.^7^ Results from the present study are consistent with Kullman et al. Such cross-sectional findings provide evidence that MetS and its clinical components are related to hypothalamic inflammation, and potentially, structural changes specific to the hypothalamus. Whether the severity of metabolic dysregulation (e.g., higher blood pressure, worse insulin resistance) or the presence of a greater number of CVD risk factors is related to the degree of inflammation and structural damage presents a focus for future investigations.

Hypothalamic gliosis is diet-induced in preclinical models.^34^ We were unable to identify consistent dietary predictors of MBH T2 signal ratios in humans in this prospective analysis. However, the prospective association between lower daily proportion of total carbohydrate consumption and greater evidence of gliosis, as assessed by the positive control T2 signal ratio, suggests that the proportions of macronutrients in daily diet might be influential. Prior findings in preclinical^35^ and human^21^ studies highlight saturated fat consumption as a possible risk factor for MBH gliosis in humans. Given the inverse relationship between carbohydrate and fat consumption, the current findings of greater evidence of gliosis with lower carbohydrate intake could be seen as supportive of the role of dietary fats in instigating hypothalamic inflammation. Furthermore, the effect of saturated fat consumption on the degree of MBH gliosis was modified by weight status. This result implies that those with overweight or obesity may have a different susceptibility to gliosis related to saturated fat consumption than individuals maintaining weight in the normal range. Further research capable of discerning risk and protective factors for hypothalamic gliosis (e.g., genetics) in relation to precisely measured dietary exposures is needed.

There are several strengths to this study. This is the largest study to date providing evidence of associations between hypothalamic gliosis and multiple CVD risk factors, including novel findings linking MBH gliosis with hypertension. Our outcomes of interest were high-quality data measured and collected by FHS investigators. Limitations to this study include the lack of statistical power to detect associations with CHD itself in a relatively young and healthy study population with few CHD cases. Additionally, participants with longstanding CHD or DM may have adapted lifestyle modification (e.g., diet changes or weight loss) with the potential to affect hypothalamic gliosis. Furthermore, most subjects self-identified as White, limiting the interpretation of our results to a select population, and we were not powered to determine sex-specific associations. MRI images are an indirect measure of MBH gliosis; higher T2 signal intensity can be due to local edema, tumors, or autoimmune inflammation, but in community-based participants in an observational study, these pathologies are unlikely. Imaging variability from factors such as B0 inhomogeneity may have limited our ability to detect smaller effect sizes. Lastly, misclassification of dietary exposures could arise due to self-report methodology and any changes in habitual intake that occurred during the several-year interim before MRI images were captured.

In sum, the current study focuses attention on the CNS and on the evolving understanding of how weight gain might relate to increased CVD risk.^36,37^ Weight gain and/or dietary factors may promote hypothalamic inflammation which acts as a shared risk factor for obesity and CVD that fails to fully resolve with lifestyle changes alone. While behavioral interventions that improve metabolic status do not always reduce CVD risk,^38^ GLP-1 agonists that promote substantial weight loss as well as bariatric surgery do benefit cardiovascular health.^39–41^ Preclinical and human studies show improvement in hypothalamic inflammation with liraglutide^42^ and bariatric surgery,^8,43^respectively.

These robust cross-sectional findings support investigations to clarify the role of hypothalamic gliosis in CVD pathogenesis. Future epidemiologic and clinical studies could examine subclinical measures of disease, such as coronary artery calcium scores and carotid intima-media thickness, to test whether hypothalamic gliosis is related to atherosclerosis prior to the manifestation of clinical disease. Additional studies should determine whether evidence of gliosis in the hypothalamus predicts incident CVD. More robust dietary assessments or integration of objective biomarkers^44^ could aid efforts to discover dietary stimulators of hypothalamic inflammation in humans. Finally, preclinical studies in animal models of atherosclerosis and HTN will be required to determine causal mechanisms whereby glial-cell mediated hypothalamic inflammation and dysfunction might drive CVD risk.

## Data Availability

All data produced in the present study are available upon reasonable request to the authors and the Framing Heart Study Administration per study guidelines.

## Non-standard Abbreviations and Acronyms

AgRP: agouti-related peptide
AMY: amygdala
BMI: body mass index
CHD: coronary heart disease
CNS: central nervous system
CVD: cardiovascular disease
DM: diabetes mellitus
FDR: false discovery rate
FFQ: food frequency questionnaire
FHS: Framingham Heart Study
HDL-C: high-density lipoprotein cholesterol
HTN: hypertension
LDL-C: low-density lipoprotein cholesterol
MBH: mediobasal hypothalamus
MetS: metabolic syndrome
MRI: magnetic resonance imaging
POMC: pro-opiomelanocortin
PUT: putamen
PVN: paraventricular nucleus
ROI: region of interest
SNA: sympathetic nerve activity
T1D: type 1 diabetes
T2D: type 2 diabetes

## Acknowledgements

From the Framingham Heart Study of the National Heart Lung and Blood Institute of the National Institutes of Health and Boston University School of Medicine. This project has been funded in whole or in part with Federal funds from the National Heart, Lung, and Blood Institute, National Institutes of Health, Department of Health and Human Services, under Contract No. 75N92019D00031. We would like to thank Yumei Feng Earley, Ph.D. for thoughtful review of the manuscript and Nash Whitford for his assistance with exponential functions.

## Supplemental Material

Figure S1-S2 Table S1–S5

