## Supplementary material for "Hypothalamic Gliosis is Associated With Multiple Cardiovascular Disease Risk Factors"

### Contents

Figures S1–S2
Tables S1–S5

Figure S1. Flow chart describing inclusion and exclusion criteria for the study population

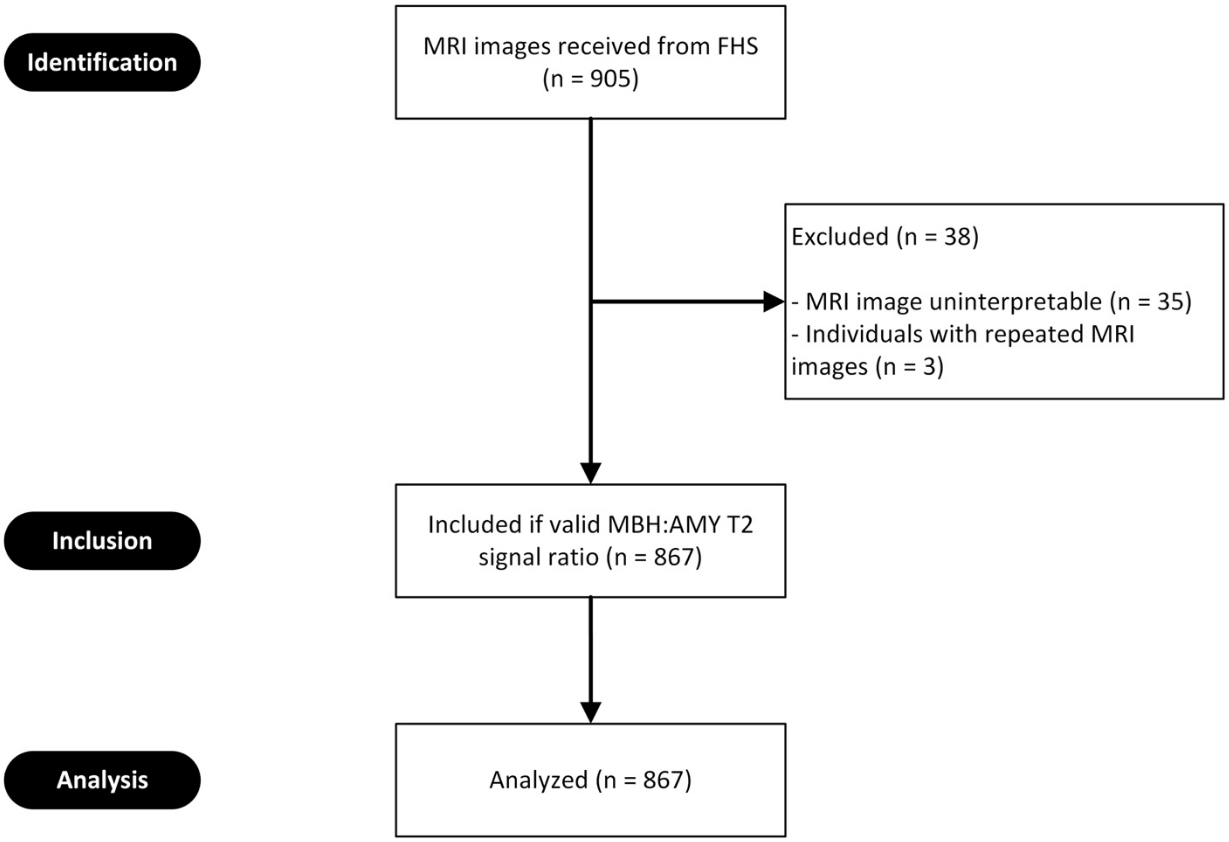


MRI, magnetic resonance imaging; FHS, Framingham Heart Study; MBH, mediobasal hypothalamus; AMY, amygdala.

Figure S2. Adjusted marginal effects model of the association of MBH/AMY T2 signal ratio and the proportion of daily saturated fat intake stratified by BMI category (<25 kg/m$^2$; ≥25 kg/m$^2$ & <30 kg/m$^2$; ≥30 kg/m$^2$)

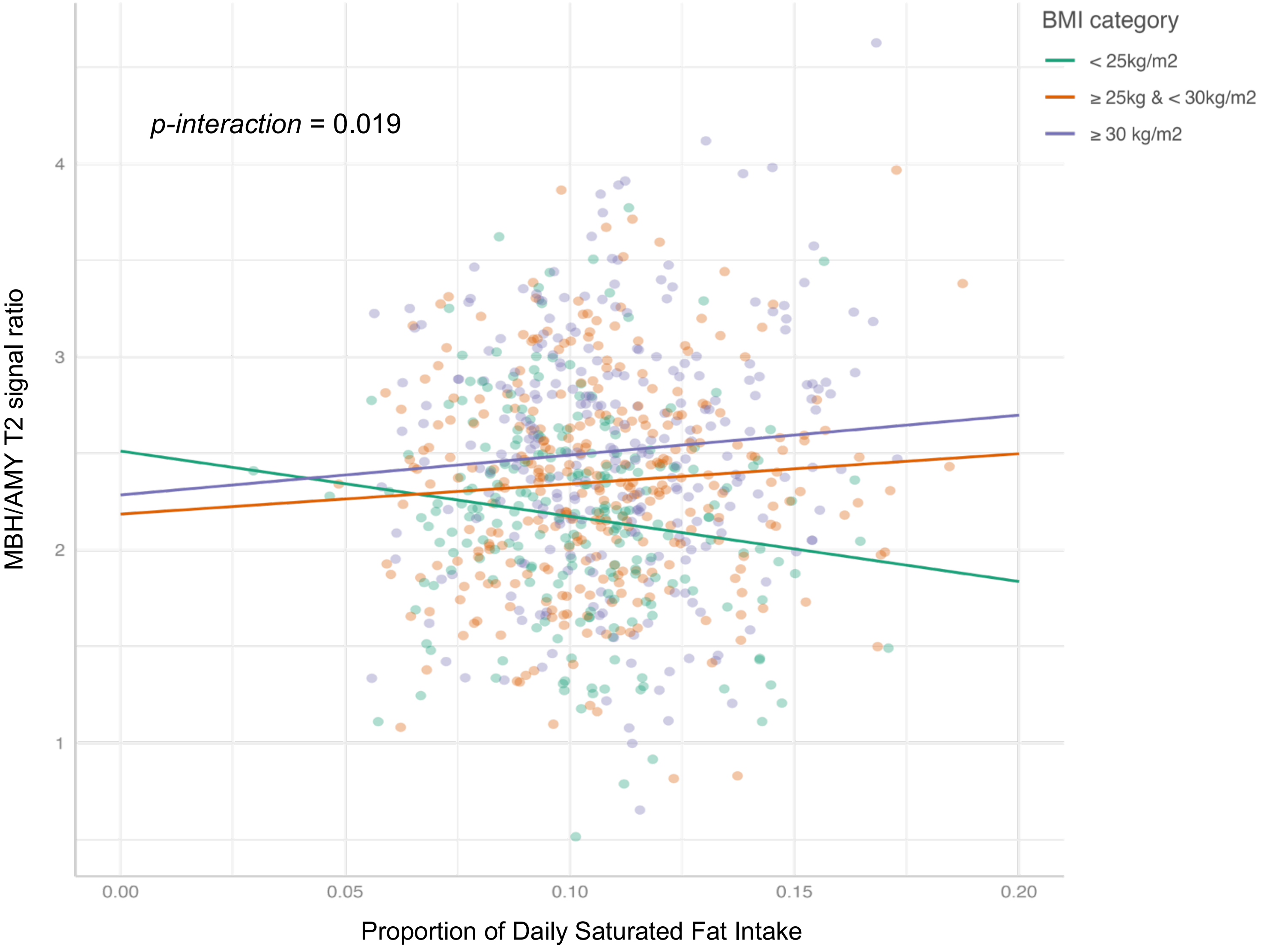


The marginal effects model was conducted using a linear regression model adjusting for age, sex, and time interval between dietary exposures and MRI and including an interaction term for BMI category*saturated fat. The sample size of each BMI category was 231, 305, and 278, respectively. MBH/AMY T2 signal ratios were natural logarithm-transformed.

Table S1. Covariates included in each model by cardiovascular risk factor and coronary heart disease outcomes.

| | BMI | HDL-C | LDL-C | ln(Triglycerides) | HTN | DM | MetS | CHD |
|---|---|---|---|---|---|---|---|---|
| Model 1 | Age, sex | Age, sex | Age, sex | Age, sex | Age, sex | Age, sex | Age, sex | Age, sex |
| Model 2 | Model 1 + Smoking | Model 1 + Smoking | Model 1 + Smoking | Model 1 + Smoking | Model 1 + Smoking | Model 1 + Smoking | Model 1 + Smoking | Model 1 + Smoking |
| Model 3 | Model 2 + Diabetes treatment | Model 2 + Lipid treatment | Model 2 + Lipid treatment | Model 2 + Lipid treatment | Model 2 + Lipid treatment | Model 2 + Hypertension | Model 2 + Physical activity | Model 2 + Lipid treatment |
| Fully adjusted | NA | Model 3 + BMI | Model 3 + BMI | Model 3 + BMI | Model 3 + BMI | Model 3 + BMI | NA | Model 3 + BMI |

BMI, body mass index; HDL-C, high-density lipoprotein cholesterol; LDL-C, low-density lipoprotein cholesterol; HTN, hypertension; DM, diabetes mellitus; MetS, metabolic syndrome; CHD, coronary heart disease

Table S2. T2 signal ratio odds ratios & 95% CIs by cardiovascular disease risk factor and coronary heart disease outcomes.

| T2 signal ratio predictor and model | | Hypertension | | Diabetes mellitus | | Metabolic syndrome | | Coronary heart disease | |
|---|---|---|---|---|---|---|---|---|---|
| | | Coefficient [95%CI] | *P* value | Coefficient [95%CI] | *P* value | Coefficient [95%CI] | *P* value | Coefficient [95%CI] | *P* value |
| MBH/AMY | Model 1* | 1.36 [1.17, 1.57] | <0.001 | 1.24 [0.95, 1.61] | 0.11 | 1.34 [1.15, 1.57] | <0.001 | 0.75 [0.49, 1.15] | 0.19 |
| | Model 2† | 1.36 [1.17, 1.57] | <0.001 | 1.24 [0.95, 1.61] | 0.12 | 1.34 [1.14, 1.57] | <0.001 | 0.73 [0.47, 1.14] | 0.17 |
| | Model 3‡ | 1.36 [1.17, 1.58] | <0.001 | 1.14 [0.87, 1.49] | 0.34 | 1.33 [1.13, 1.56] | <0.001 | 0.71 [0.45, 1.12] | 0.14 |
| | Fully adjusted§ | 1.23 [1.05, 1.44] | 0.0088 | 1.01 [0.77, 1.33] | 0.93 | … | … | 0.69 [0.44, 1.08] | 0.11 |
| MBH/PUT | Model 1* | 1.24 [1.08, 1.43] | 0.0030 | 1.39 [1.07, 1.81] | 0.015 | 1.35 [1.15, 1.58] | <0.001 | 0.99 [0.67, 1.47] | 0.96 |
| | Model 2† | 1.25 [1.08, 1.44] | 0.0028 | 1.37 [1.06, 1.79] | 0.018 | 1.34 [1.15, 1.57] | <0.001 | 0.97 [0.65, 1.45] | 0.89 |
| | Model 3‡ | 1.24 [1.07, 1.43] | 0.0040 | 1.30 [1.00, 1.69] | 0.051 | 1.34 [1.14, 1.57] | <0.001 | 0.97 [0.65, 1.44] | 0.87 |
| | Fully adjusted§ | 1.13 [0.97, 1.31] | 0.12 | 1.19 [0.91, 1.56] | 0.21 | … | … | 0.95 [0.64, 1.42] | 0.81 |
| PUT/AMY | Model 1* | 0.99 [0.86, 1.14] | 0.88 | 0.81 [0.62, 1.05] | 0.11 | 0.90 [0.77, 1.05] | 0.18 | 0.82 [0.55, 1.23] | 0.35 |
| | Model 2† | 0.99 [0.86, 1.14] | 0.85 | 0.82 [0.63, 1.06] | 0.12 | 0.91 [0.78, 1.05] | 0.20 | 0.85 [0.57, 1.26] | 0.41 |
| | Model 3‡ | 1.00 [0.86, 1.15] | 0.95 | 0.81 [0.63, 1.05] | 0.12 | 0.90 [0.78, 1.05] | 0.20 | 0.84 [0.56, 1.25] | 0.39 |
| | Fully adjusted§ | 1.02 [0.88, 1.18] | 0.83 | 0.83 [0.63, 1.09] | 0.18 | … | … | 0.83 [0.55, 1.25] | 0.37 |

Results are from multiple logistic regression models. MRI-assessed T2 signal ratios were natural logarithm transformed and used as model predictors: MBH/AMY (primary), MBH/PUT (positive control), and PUT/AMY (negative control). ORs and 95% CIs are presented as the change in odds for the outcome per a 1 SD difference in T2 signal ratio.

MBH, mediobasal hypothalamus; AMY, amygdala; PUT, putamen; OR, odds ratio, CI, confidence interval.

* Model 1 adjusted for age and sex.
† Model 2 adjusted for model 1 covariates plus smoking.
‡ Model 3 adjusted for model 2 covariates plus lipid treatment for hypertension and CHD models, hypertension status for diabetes mellitus models, or physical activity index for metabolic syndrome models.
§ Fully adjusted model includes model 3 covariates plus BMI.

Table S3. T2 signal ratio odds ratios & 95% CIs by continuous cardiovascular risk factor outcomes, adjusting for time between MRI assessment and health exam.

| T2 signal ratio predictor and model | | BMI | | HDL-C | | LDL-C | | Ln(Triglycerides) | |
|---|---|---|---|---|---|---|---|---|---|
| | | Coefficient [95%CI] | *P* value | Coefficient [95%CI] | *P* value | Coefficient [95%CI] | *P* value | Coefficient [95%CI] | *P* value |
| MBH/AMY | Model 1* | 22.1 [15.9, 28.2] | <0.001 | -47.1 [-67.3, -27.0] | <0.001 | 18.2 [-16.5, 52.8] | 0.30 | 1.2 [0.6, 1.7] | <0.001 |
| | Model 2† | 22.1 [15.9, 28.2] | <0.001 | -47.1 [-67.3, -26.9] | <0.001 | 17.7 [-16.9, 52.4] | 0.32 | 1.1 [0.6, 1.7] | <0.001 |
| | Model 3‡ | 21.4 [15.4, 27.5] | <0.001 | -46.1 [-66.1, -26.1] | <0.001 | 21.5 [-11.4, 54.4] | 0.20 | 1.1 [0.6, 1.7] | <0.001 |
| | Fully adjusted§ | … | … | -20.8 [-40.0, -1.6] | 0.034 | 12.5 [-21.2, 46.3] | 0.47 | 0.5 [-0.0, 1.0] | 0.076 |
| MBH/PUT | Model 1* | 13.0 [8.6, 17.3] | <0.001 | -34.7 [-48.8, -20.6] | <0.001 | -1.8 [-25.9, 22.2] | 0.88 | 0.9 [0.5, 1.3] | <0.001 |
| | Model 2† | 13.2 [8.9, 17.5] | <0.001 | -34.1 [-48.2, -20.0] | <0.001 | -1.0 [-25.1, 23.1] | 0.94 | 0.9 [0.5, 1.2] | <0.001 |
| | Model 3‡ | 12.5 [8.2, 16.8] | <0.001 | -32.8 [-46.7, -18.8] | <0.001 | 3.3 [-19.6, 26.3] | 0.78 | 0.8 [0.5, 1.2] | <0.001 |
| | Fully adjusted§ | … | … | -18.1 [-31.4, -4.8] | 0.0079 | -2.8 [-26.1, 20.5] | 0.81 | 0.5 [0.1, 0.8] | 0.012 |
| PUT/AMY | Model 1* | -2.8 [-7.7, 2.1] | 0.26 | 15.0 [-0.8, 30.8] | 0.063 | 17.3 [-9.4, 44.0] | 0.20 | -0.4 [-0.8, 0.0] | 0.077 |
| | Model 2† | -3.1 [-8.0, 1.8] | 0.22 | 14.2 [-1.7, 30.0] | 0.080 | 16.1 [-10.7, 42.8] | 0.24 | -0.4 [-0.8, 0.1] | 0.10 |
| | Model 3‡ | -2.6 [-7.4, 2.3] | 0.30 | 12.9 [-2.8, 28.6] | 0.11 | 12.4 [-13.1, 37.9] | 0.34 | -0.3 [-0.8, 0.1] | 0.12 |
| | Fully adjusted§ | … | … | 9.8 [-4.7, 24.4] | 0.19 | 13.5 [-11.9, 38.9] | 0.30 | -0.3 [-0.7, 0.1] | 0.20 |

Results are from multiple linear regression models. MRI-assessed T2 signal ratios were natural logarithm transformed and used as model predictors: MBH/AMY (primary), MBH/PUT (positive control), and PUT/AMY (negative control). Coefficient and confidence intervals represent the estimated change in outcome per 1 unit difference in the natural logarithm-transformed T2 signal ratio.

BMI, body mass index; HDL-C, high-density lipoprotein cholesterol; LDL-C, low-density lipoprotein cholesterol; ln(Triglycerides), natural logarithm transformed fasting triglycerides; MBH, mediobasal hypothalamus; AMY, amygdala; PUT, putamen.

* Model 1 adjusted for age, sex, and interval between health examination and MRI assessment.
† Model 2 adjusted for model 1 covariates plus smoking.
‡ Model 3 adjusted for model 2 covariates plus diabetes treatment for BMI model or lipid treatment for HDL-C, LDL-C, and natural log-transformed triglycerides models.
§ Fully adjusted model includes model 3 covariates plus BMI, when appropriate.

Table S4. T2 signal ratio odds ratios & 95% CIs by cardiovascular risk factor and coronary heart disease outcomes, adjusting for time between MRI assessment and health exam.

| T2 signal ratio predictor and model | | Hypertension | | Diabetes mellitus | | Metabolic syndrome | | Coronary heart disease | |
|---|---|---|---|---|---|---|---|---|---|
| | | Coefficient [95%CI] | *P* value | Coefficient [95%CI] | *P* value | Coefficient [95%CI] | *P* value | Coefficient [95%CI] | *P* value |
| MBH/AMY | Model 1* | 1.4 [1.2, 1.6] | <0.001 | 1.2 [0.9, 1.6] | 0.12 | 1.3 [1.1, 1.6] | <0.001 | 0.7 [0.5, 1.1] | 0.18 |
| | Model 2† | 1.4 [1.2, 1.6] | <0.001 | 1.2 [0.9, 1.6] | 0.12 | 1.3 [1.1, 1.6] | <0.001 | 0.7 [0.5, 1.1] | 0.17 |
| | Model 3‡ | 1.4 [1.2, 1.6] | <0.001 | 1.1 [0.9, 1.5] | 0.34 | 1.3 [1.1, 1.6] | <0.001 | 0.7 [0.4, 1.1] | 0.10 |
| | Fully adjusted§ | 1.2 [1.1, 1.4] | 0.0088 | 1.0 [0.8, 1.3] | 0.94 | … | … | 0.7 [0.4, 1.1] | 0.081 |
| MBH/PUT | Model 1* | 1.2 [1.1, 1.4] | 0.0027 | 1.4 [1.1, 1.8] | 0.011 | 1.4 [1.2, 1.6] | <0.001 | 1.0 [0.7, 1.4] | 0.88 |
| | Model 2† | 1.2 [1.1, 1.4] | 0.0024 | 1.4 [1.1, 1.8] | 0.013 | 1.4 [1.2, 1.6] | <0.001 | 1.0 [0.6, 1.4] | 0.82 |
| | Model 3‡ | 1.2 [1.1, 1.4] | 0.0036 | 1.3 [1.0, 1.7] | 0.043 | 1.3 [1.1, 1.6] | <0.001 | 0.9 [0.6, 1.4] | 0.72 |
| | Fully adjusted§ | 1.1 [1.0, 1.3] | 0.11 | 1.2 [0.9, 1.6] | 0.20 | … | … | 0.9 [0.6, 1.4] | 0.68 |
| PUT/AMY | Model 1* | 1.0 [0.9, 1.1] | 0.85 | 0.8 [0.6, 1.0] | 0.087 | 0.9 [0.8, 1.0] | 0.15 | 0.8 [0.6, 1.2] | 0.37 |
| | Model 2† | 1.0 [0.9, 1.1] | 0.82 | 0.8 [0.6, 1.0] | 0.10 | 0.9 [0.8, 1.0] | 0.17 | 0.9 [0.6, 1.3] | 0.43 |
| | Model 3‡ | 1.0 [0.9, 1.1] | 0.92 | 0.8 [0.6, 1.0] | 0.11 | 0.9 [0.8, 1.0] | 0.17 | 0.8 [0.6, 1.3] | 0.41 |
| | Fully adjusted§ | 1.0 [0.9, 1.2] | 0.83 | 0.8 [0.6, 1.1] | 0.17 | … | … | 0.8 [0.6, 1.3] | 0.39 |

Results are from multiple logistic regression models. MRI-assessed T2 signal ratios were natural logarithm transformed and used as model predictors: MBH/AMY (primary), MBH/PUT (positive control), and PUT/AMY (negative control). ORs and 95% CIs are presented as the change in odds for the outcome per a 1 SD difference in T2 signal ratio.

MBH, mediobasal hypothalamus; AMY, amygdala; PUT, putamen; OR, odds ratio, CI, confidence interval.

* Model 1 adjusted for age, sex, and interval between health examination and MRI assessment.
† Model 2 adjusted for model 1 covariates plus smoking.
‡ Model 3 adjusted for model 2 covariates plus lipid treatment for hypertension and CHD models, hypertension status for diabetes mellitus models, or physical activity index for metabolic syndrome models.
§ Fully adjusted model includes model 3 covariates plus BMI.

Table S5. Adjusted prospective associations of self-reported dietary exposures and T2 signal ratio outcomes (N=814).

| | MBH/AMY T2 signal ratio | | MBH/PUT T2 signal ratio | | PUT/AMY T2 signal ratio | |
|---|---|---|---|---|---|---|
| Predictor* | Coefficient [95% CI] | *P* value | Coefficient [95% CI] | *P* value | Coefficient [95% CI] | *P* value |
| Total fat | 0.04 [-0.02, 0.11] | 0.19 | 0.03 [-0.07, 0.13] | 0.57 | 0.01 [-0.07, 0.10] | 0.76 |
| Total carbohydrates | -0.04 [-0.09, 0.01] | 0.15 | -0.08 [-0.16, 0.00] | 0.04 | 0.04 [-0.03, 0.11] | 0.25 |
| Total protein | 0.09 [-0.04, 0.22] | 0.19 | 0.16 [-0.03, 0.35] | 0.10 | -0.06 [-0.23, 0.11] | 0.50 |
| Fructose | -0.03 [-0.22, 0.17] | 0.78 | -0.02 [-0.30, 0.26] | 0.89 | -0.01 [-0.27, 0.24] | 0.93 |
| Sucrose | -0.08 [-0.20, 0.04] | 0.19 | -0.2 [-0.38, -0.02] | 0.025 | 0.10 [-0.06, 0.27] | 0.21 |
| Total saturated fat | 0.12 [-0.04, 0.29] | 0.14 | 0.18 [-0.07, 0.42] | 0.15 | -0.05 [-0.27, 0.17] | 0.63 |
| Total sugar | -0.02 [-0.08, 0.05] | 0.63 | -0.04 [-0.13, 0.05] | 0.44 | 0.02 [-0.07, 0.10] | 0.70 |

T2 signal ratios were natural log-transformed. All models were adjusted for age, sex, and time interval between dietary exposures and MRI.

* Predictors were calculated as proportions of total caloric intake.
